# Incorporating Stroke Severity Dynamics to Improve Prognostic Modelling in Ischaemic Stroke Patients: Evidence from Serial NIHSS Assessments in Bergen NORSTROKE Study

**DOI:** 10.1101/2025.11.08.25339813

**Authors:** Z Lu, H Næss, M Gittins, AK Kishore, CJ Smith, A Vail

## Abstract

**Background:** Stroke severity evolves rapidly in the acute phase, and yet severity is commonly measured only on admission and treated as a static, single-timepoint measure in stroke research. This study explores routinely collected, repeated National Institute of Health Stroke Scale (NIHSS) assessments to better understand the dynamic nature of stroke severity and evaluate its value for outcome prediction.

**Methods:** We analysed real-world data from 1,622 ischaemic stroke patients admitted to the Bergen NORSTROKE study, all of whom had at least two NIHSS assessments within 48 hours of symptom onset. Stroke severity trajectories were explored using summary statistics, spaghetti plots, linear mixed-effects models (LMM), and group-based trajectory modelling (GBTM). Seven logistic regression models were compared for their ability to predict favourable short-term functional outcomes (modified Rankin Scale), incorporating various representations of stroke severity dynamics. Model performance was evaluated using AIC, BIC, and area under the ROC curve (AUC).

**Results:** NIHSS scores varied considerably over time. Most patients had mild strokes (median NIHSS = 2) and showed improvement within 48 hours. GBTM identified three latent groups: Very Low-Stable (40.3%), Moderate Low-Stable (41%), and High-Mildly Improving (18.7%). Models incorporating stroke severity dynamics outperformed those using admission NIHSS alone (AUC 0.835 vs. 0.778). The best predictive performance was achieved using random intercepts and slopes from the LMM. While GBTM improved model fit (AIC = 1624), its added discriminatory power was limited (AUC = 0.792).

**Conclusion:** Stroke severity evolves dynamically in the acute phase. Incorporating these dynamics into prognostic models improves predictive accuracy and model fit. Advanced modelling approaches that account for individual symptom trajectories offer a more accurate and clinically relevant framework for stroke outcome prediction. This also underscores the importance of incrementally updating prediction models with new clinical information, as changing severity is a key but often overlooked element in stroke studies.

## Introduction

Stroke is a leading cause of death and long-term disability worldwide, with ischaemic stroke accounting for the vast majority of cases ^1, 2^. The clinical presentation of stroke is dynamic, especially in the acute phase following symptom onset ^3^. For ischaemic stroke patients, neurological symptoms can evolve rapidly due to ongoing brain injury and reperfusion ^4, 5^. Understanding these dynamics is important to inform acute stroke care management and improve stroke prognostication.

The National Institutes of Health Stroke Scale (NIHSS) is a standardised tool widely used to assess the severity of stroke ^6, 7^. While routinely collected at hospital admission, NIHSS is typically measured only once and used as a static marker of baseline stroke severity ^8^. In many prognostic studies, admission NIHSS is treated as a confounder or a primary predictor without consideration of its potential variability over time ^9, 10, 11^.

However, our previous research highlights a dynamic interplay between stroke severity and time to assessment ^12, 13^. Patients with more severe symptoms tend to present earlier, resulting in a shorter time between symptom onset and NIHSS assessment ^14^. Conversely, time to assessment itself can influence the severity score, as symptoms may improve or worsen within hours. This bidirectional relationship means that stroke severity is not fixed but evolves over time, and incorporating these dynamics into prognostic models may enhance their predictive accuracy and clinical relevance.

There is increasing interest in modelling stroke severity trajectories to capture symptom progression and improve both prediction and explanation of outcomes. Studies using clinical trial data have shown that trajectory-based models often outperform single-timepoint approaches. However, such studies typically rely on data collected at fixed time points (e.g., pre-randomisation, post-treatment, days 2, 3, 5, 7), limiting their generalisability. In contrast, routine clinical data are more complex characterised by irregular time intervals, missing values, and variable numbers of assessments across patients. Importantly, the frequency of measurement itself may depend on both baseline severity and observed changes in severity, adding further challenges to analysis but also offering a more realistic basis for model development and validation.

Nevertheless, most existing prognostic models rely on fixed admission measures, partly because they are simple, widely available, and routinely collected. This approach has clear limitations, which has led to growing attention on dynamic prediction models and causal prediction frameworks designed to improve the reliability and interpretability of prognostic modelling.^15^.

This study aims to address this gap by leveraging real-world, routinely collected data to investigate the dynamic nature of stroke severity. Specifically, we assess whether NIHSS scores change meaningfully over time after symptom onset, and if so, how these changes manifest across patients. We then evaluate whether incorporating the temporal dynamics of NIHSS improves stroke outcome prediction by comparing a range of modelling approaches for repeated measurements and examining their added prognostic value.

## Methods

### Study design and settings

We undertook a secondary analysis of data initially collected for the Bergen NORSTROKE study. Patients were admitted to a single Stroke Unit, Department of Neurology, Haukeland University Hospital, between February 2006 and February 2021, which routinely assessed NIHSS repeatedly at irregular intervals for up to 14 times in the first 2 days post admission for patient monitoring purposes.

### Participants and eligibility

Patients with an ischaemic stroke were included. Ischaemic stroke is defined as an episode of neurological dysfunction caused by focal cerebral, spinal, or retinal infarction confirmed in computer tomography (CT) or magnetic resonance imaging (MRI).

We censored the NIHSS assessment after hyperacute treatment including intravenous thrombolysis and mechanical thrombectomy to examine the natural dynamics unaffected by known treatments. We included patients who have at least two NIHSS assessments within two days since symptom onset to be able to evaluate and model changes in NIHSS.

We excluded patients with transient ischaemic attacks (TIA), brain haemorrhage or unknown stroke types. Patients with missing time of assessments or wrong recorded time were excluded from the study. We also excluded patients with missing outcomes in prognostic models.

### Statistical analysis

We used summary statistics to explore patterns of change in stroke severity over time. Based on individual-level NIHSS change scores, we classified patients into five categories: (1) *Improved*—all change scores are less than or equal to zero; (2) *Deteriorated*—all change scores are non-negative, with at least one positive; (3) *Mixed-Deteriorated*—both negative and positive changes are present, and the first non-zero change is positive; (4) *Mixed-Improved*—both negative and positive changes are present, and the first non-zero change is negative; and (5) *Unchanged*—all change scores are zero. To visualise individual trajectories of stroke severity, we created spaghetti plots for each category, with the deteriorated and unchanged groups combined because of their small proportions and to improve clarity of presentation.

We employed linear mixed-effects models (LMM) to analyse the repeated NIHSS measurements, capturing within-patient variation through fixed effects and between-patient variation through random effects ^16, 17, 18^. Both time from symptom onset to assessment (in hour unit) and stroke severity (NIHSS) were treated as continuous variables. We fitted models with random intercepts and random slopes to account for individual-specific baseline severity and rates of change. To model potential non-linear relationships between time and stroke severity, we also incorporated polynomial terms. Model performance was compared using Akaike Information Criterion (AIC) and Bayesian Information Criterion (BIC) to identify the best-fitting approach ^19^.

We also used group-based trajectory modelling (GBTM) to further identify three pre-specified latent subgroups with distinct trajectories of stroke severity. In this context, “pre-specified” refers to setting the number of subgroups in advance, based on prior knowledge, theoretical expectations, or model fit criteria, rather than letting the data alone dictate an arbitrary number of groups. This differs from the naïve classification approach, where patients were categorised directly from their individual-level NIHSS change scores. In contrast, GBTM estimates latent subgroups probabilistically and models longitudinal data over time.

GBTM is a flexible approach used to identify unobserved subgroups of individuals who share similar longitudinal patterns ^20, 21^. It estimates each individual’s probability of belonging to different trajectory groups and fits separate models within each group, typically using polynomial functions of time to capture changes in stroke severity. Group membership probabilities are modelled via multinomial logistic regression with only intercepts. By assuming that within-person variation is fully explained by group trajectories, GBTM treats repeated measures as independent within each group. The model outputs group-specific trajectories and individual-level posterior probabilities of group membership.

To assess the prognostic utility of different ways of incorporating stroke severity dynamics, we compared the performance of seven logistic regression models predicting favourable functional outcome at day seven (or at discharge if earlier), defined as a dichotomised modified Rankin Scale (mRS). These models can be grouped into three sets, reflecting different practical scenarios in which prediction might occur. The first set focused on information available at admission: (1) admission NIHSS only (benchmark); (2) admission NIHSS, time from symptom onset to assessment, and their interaction (based on our previous study). The second set updated predictions once a second NIHSS assessment became available: (3) admission NIHSS and second NIHSS measurement; (4) model (2) with the second NIHSS added. The third set used information on trajectories up to seven days (or discharge if earlier): (5) group-based trajectory modelling (GBTM)–derived latent class as a categorical predictor; (6) a naïve five-level classification based on individual change scores; (7) individual-level random intercepts and slopes from the best-fitting linear mixed-effects model as continuous predictors.

This structure highlights that models (1) to (2), (3) to (4), and (5) to (7) represent three stages of a dynamic updating process, consistent with modern approaches to prediction modelling. Each set has a different underlying aim depending on the timing of the available data: initial prognostication at admission, updated prediction after further NIHSS assessment, and trajectory-informed prediction over the first two days of care.

Model performance was evaluated using AIC and BIC for model fit, and the area under the ROC curve (AUC) to assess discrimination ability ^22^.

## Results

The flowchart (Figure 1) illustrates the process of data cleaning and patient selection for the study. Out of 2686 initially identified patients with at least two NIHSS measurements, 514 were excluded during the data cleaning stage due to missing treatment information (n = 358), or incorrect time recordings (n = 261). This left 2172 patients for eligibility screening. A further 550 patients were excluded due to having a diagnosis of transient ischaemic attack (n = 314), brain haemorrhage, ICH specifically (n = 234), or unknown stroke types (n = 2), resulting in 1622 patients eligible for trajectory modelling in stage one. One patient was excluded due to missing outcome data, leaving a final sample of 1621 patients for prognostic modelling in stage two.

**Figure 1.**
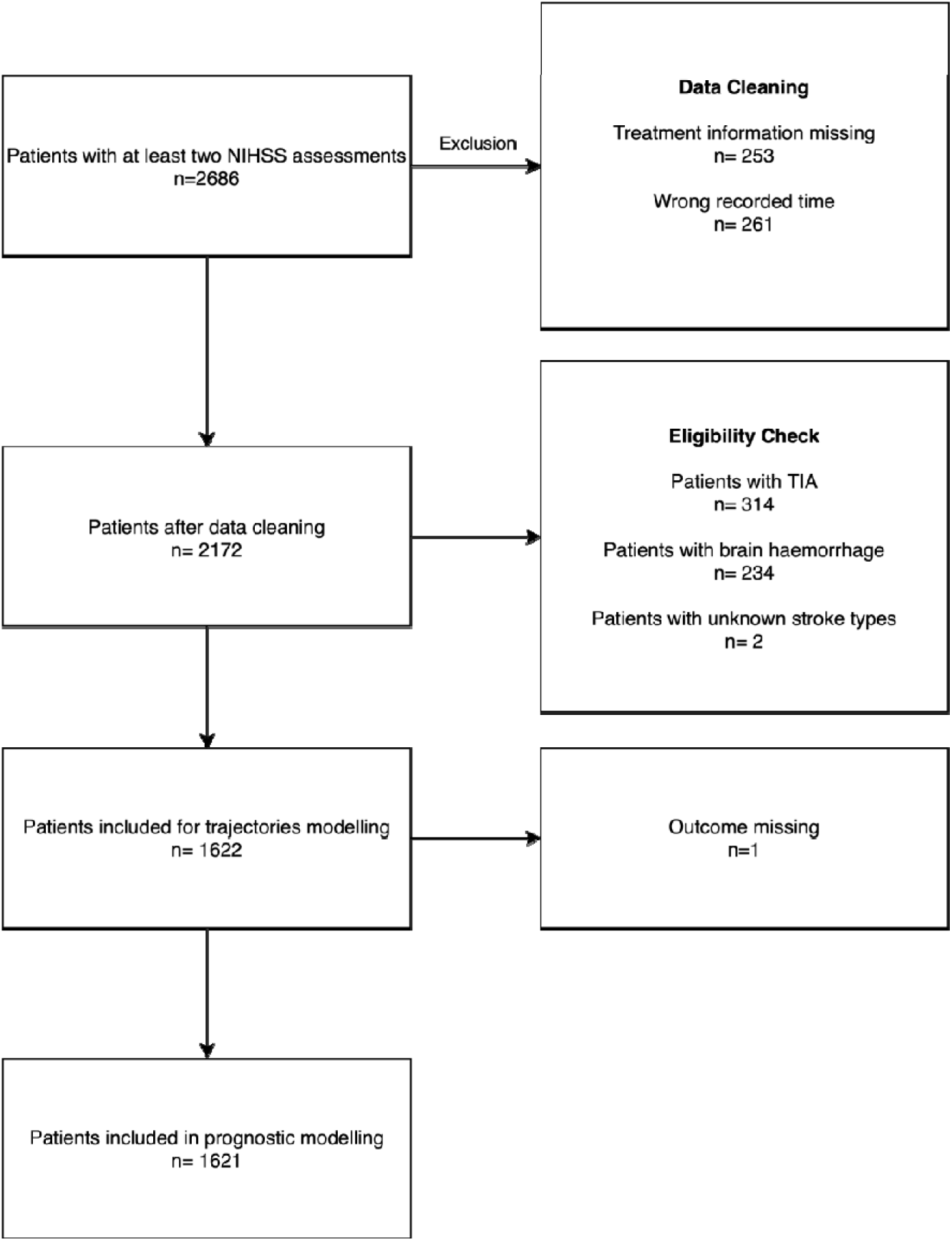
Flow chart of data cleaning and patient selection

Table 1 summarises the characteristics of the study cohort comprising 1,622 patients. The median age was 74 years (interquartile range [IQR]: 62–83), and 42% of the patients were male. The median time from symptom onset to the first NIHSS assessment was 5 hours (IQR: 2–13), while the median time to the second assessment was 8 hours (IQR: 4–17). The median NIHSS score at admission was 2 (IQR: 1–5) and remained similar at the second measurement (median: 2; IQR: 0–5). In terms of changes in stroke severity over time, 27.7% of patients showed improvement, 34% had mixed-deteriorated patterns, 23% had mixed-improved patterns, 15% remained unchanged, and only 0.3% experienced clear deterioration.

**Table 1.** Patient characteristics.

| **Characteristics** | **Study cohort**<br>**(n=1622)** |
| --- | --- |
| Demographic |  |
| Age, median (IQR) y | 74 (62 to 83) |
| Sex, (Male=1) % | 42 |
| Time from symptom onset to admission NIHSS<br>assessment, median (IQR) hrs | 5 (2 to 13) |
| Time from symptom onset to the second NIHSS<br>assessment, median (IQR) hrs | 8 (4 to 17) |
| Admission NIHSS, median (IQR) | 2 (1 to 5) |
| Second NIHSS, median (IQR) | 2 (0 to 5) |
| Summary of changes in stroke severity, n (%) |  |
| Deteriorated | 5 (0.3) |
| Improved | 449 (27.7) |
| Mixed- Deteriorated | 545 (34) |
| Mixed- Improved | 370 (23) |
| Unchanged | 253 (15) |

The spaghetti plots (Figure 2) display individual NIHSS trajectories stratified by patient categories, highlighting substantial heterogeneity in stroke severity progression. The combined deteriorated/unchanged group shows relatively flat or stable patterns. In contrast, the mixed-improved and mixed-deteriorated groups demonstrate fluctuating courses, with both gains and losses in neurological function over time. The improved group predominantly shows gradual recovery, whereas the mixed-deteriorated group includes several patients with worsening severity. Together, these patterns underscore the diverse and dynamic nature of stroke progression across patients. This visual pattern aligns with the summary statistics in Table 1, where only 0.3% of patients showed clear deterioration, 27.7% improved, and over half (57%) had mixed trajectories—either initial improvement followed by deterioration or vice versa—highlighting the dynamic and non-linear nature of stroke recovery in the acute phase.

**Figure 2.**
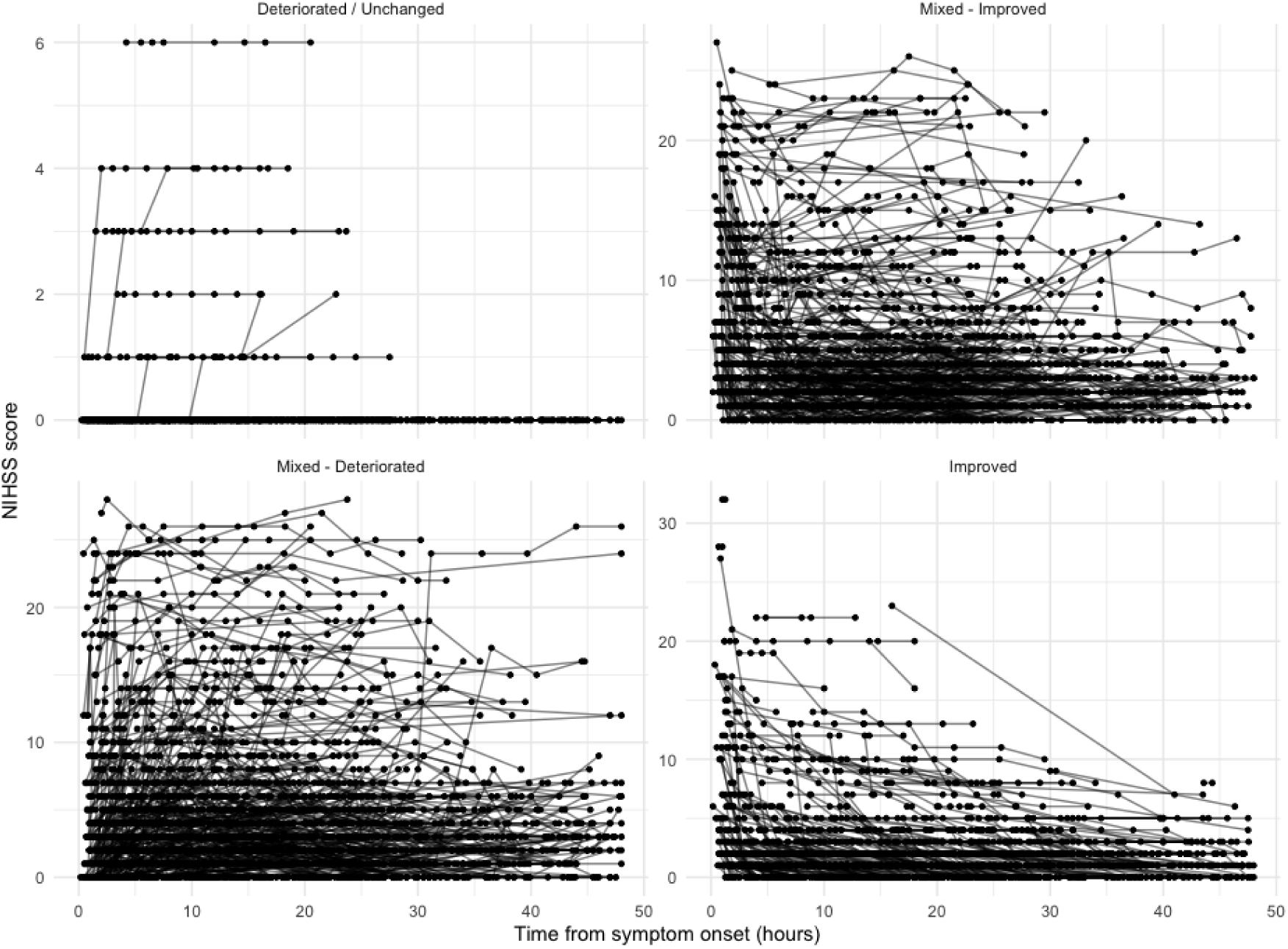
The spaghetti plots to visualise stroke severity trajectories for all patients in four naïve categories: Deteriorated and unchanged combined; Mixed improved; Mixed deteriorated; Improved

The results of model selection for the linear mixed-effects models (Table 2) show that incorporating both random intercepts and random slopes improved model fit compared with random intercept-only models, as indicated by lower AIC and BIC values across both linear and quadratic specifications. Between the random slope models, the linear specification provided the best fit (AIC = 36011, BIC = 36053), slightly outperforming the quadratic polynomial model (AIC = 36024, BIC = 36073). This suggests that allowing patient-specific trajectories with a simple linear time effect was sufficient to capture the observed variation in NIHSS scores, with limited additional benefit from including a quadratic term.

**Table 2.**
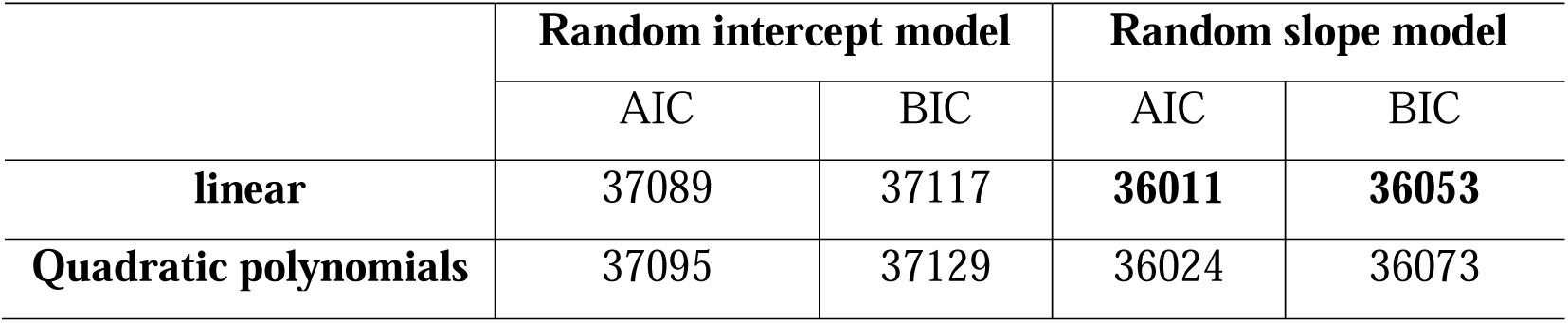
Linear mixed effect model selection.

|  | Random intercept model |  | Random slope model |  |
| --- | --- | --- | --- | --- |
|  | AIC | BIC | AIC | BIC |
| **linear** | 37089 | 37117 | **36011** | **36053** |
| **Quadratic polynomials** | 37095 | 37129 | 36024 | 36073 |

Tables 3 and 4 present the parameter estimates of the selected linear random slope model. The fixed effects indicate a modest but statistically significant negative trend over time (Estimate = −0.03, t = −7.3), suggesting that, on average, NIHSS scores decrease slightly as time progresses from symptom onset. Substantial between-patient variability is observed in baseline stroke severity (random intercept SD = 4.9) and in individual rates of change over time (random slope SD = 0.1), with a moderate negative correlation (−0.4), indicating that patients with higher initial severity tend to improve more slowly. The residual standard deviation (1.3) captures within-person fluctuations in severity not explained by the model.

**Table 3.** Fixed effect estimates for the linear random slope model.

| Fixed effects | Estimates | Standard error | t values |
| --- | --- | --- | --- |
| **Intercept** | 4.0 | 0.1 | 30.4 |
| **Time** | -0.03 | 0.004 | -7.3 |

**Table 4.**
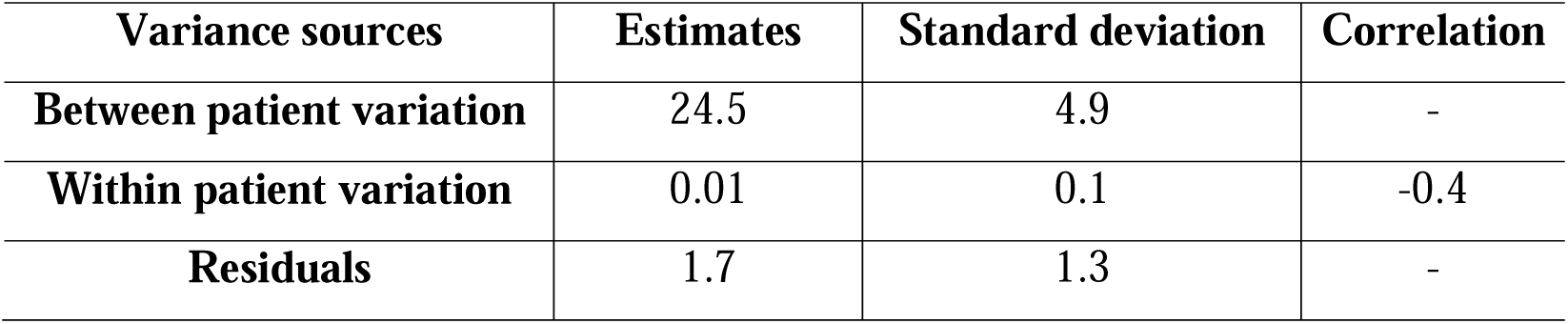
Variance estimates for the linear random slope model.

GBTM, with the number of classes pre-specified as three, assigned patients into latent subgroups based on the evolution of their NIHSS scores within 48 hours of symptom onset. Classification was probabilistic, with most patients having a dominant posterior probability for one class, though some showed more evenly distributed probabilities across classes. As shown in Table 5, the model classified 654 patients (40.3%) into Class 1, 303 patients (18.7%) into Class 2, and 665 patients (41%) into Class 3. The trajectories for these classes are illustrated in Figure 3. Class 1, labelled as the “Very Low-Stable” group, comprises patients with consistently low NIHSS scores over time. Class 2, the “High-Mildly Improving” group, represents patients with initially high NIHSS scores followed by a slight decrease. Class 3, termed the “Moderate Low-Stable” group, includes individuals with intermediate NIHSS scores that remain relatively constant. These distinct patterns reflect heterogeneous recovery profiles and provide insights into the dynamic nature of early stroke severity.

**Figure 3.**
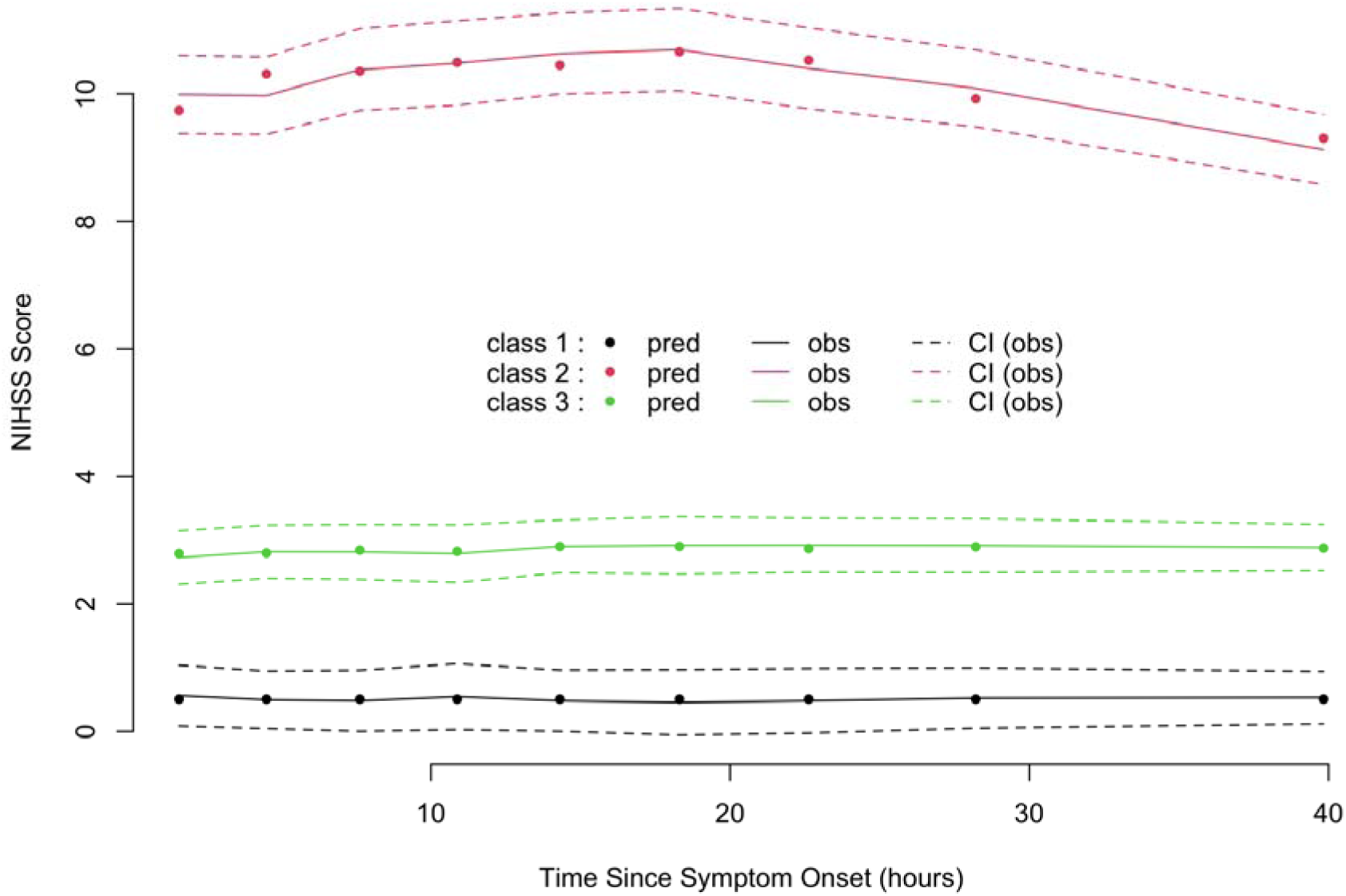
Latent groups identified by group-based trajectory modelling. Solid lines represent the predicted mean trajectory for each latent group (class 1 in black, class 2 in red, and class 3 in green). Dots connected by dashed lines indicate the observed group mean trajectories based on the data. The surrounding dashed curves represent the 95% confidence intervals for the observed means, illustrating the degree of uncertainty around each group’s estimated trajectory.

**Table 5.** Posterior classification of individuals based on group-based trajectory modelling.

|  | **Class1**<br>**Very Low-Stable** | **Class2**<br>**High-Mildly**<br>**Improving** | **Class3**<br>**Moderate Low-**<br>**Stable** | **Total** |
| --- | --- | --- | --- | --- |
| **n** | 654 | 303 | 665 | 1622 |
| **%** | 40.3 | 18.7 | 41 | 100 |

The performance comparison of seven prognostic models summarised in Table 6 highlights the importance of capturing stroke severity dynamics beyond a single baseline measurement and incrementally updating the prediction model. Using only the admission NIHSS (Model 1) provided modest discrimination (AUC = 0.778). Incorporating time and its interaction with admission NIHSS (Model 2) slightly improved both model fit and discrimination (AUC = 0.780), but the gain was marginal. Improvements were observed when a second NIHSS measurement was included (Models 3 and 4), reflecting the prognostic value of tracking severity changes over time. The GBTM-derived latent classes (Model 5) markedly improved model fit (lowest AIC/BIC among categorical models), but the increase in AUC (0.792) was modest, suggesting that while GBTM captures meaningful trajectory patterns, its categorical nature may limit discriminatory precision. In contrast, the naïve five-category change classification (Model 6) performed poorly (AUC = 0.682), indicating that summary-based categorisation fails to sufficiently capture stroke dynamics for prediction. Finally, Model 7, which used individual-specific random intercepts and slopes from the best LMM, achieved the highest performance (AUC = 0.835), demonstrating that modelling individual variation in both baseline severity and change over time is most effective. Overall, these findings underscore that incorporating dynamic stroke severity using formal modelling approaches— rather than crude summary statistics or single static stroke severity—improves prognostic model accuracy and efficiency.

**Table 6.** Comparison of prognostic model performance.

| Models | Performance matrix |  |  |
| --- | --- | --- | --- |
|  | AIC | BIC | AUC |
| 1- Admission NIHSS only | 1759 | 1770 | 0.778 |
| 2- Admission NIHSS + time + interaction | 1750 | 1772 | 0.780 |
| 3- Admission NIHSS + second NIHSS | 1657 | 1637 | 0.805 |
| 4- Admission NIHSS + time + interaction + second NIHSS | 1654 | 1681 | 0.808 |
| 5- GBTM latent category | 1624 | 1640 | 0.792 |
| 6- Crude classification based on NIHSS change scores | 1921 | 1948 | 0.682 |
| 7- LMM intercepts and slopes | 1504 | 1520 | 0.835 |

## Discussion

This study demonstrates that stroke severity, as measured by NIHSS, evolves dynamically over time following symptom onset. Our analyses, including both naïve classifications and advanced linear mixed-effects modelling, support the view that symptom severity is not static. Group-based trajectory modelling further revealed three distinct longitudinal patterns of severity. The majority of patients in our cohort experienced mild strokes and showed early improvement within the first 48 hours. Importantly, incorporating stroke severity dynamics into prognostic models, rather than relying solely on admission NIHSS or simplified change summaries, improved model fit and predictive performance.

Our findings align with prior research supporting the dynamic nature of stroke severity. Three published studies using GBTM identified three trajectory groups, including a severe symptom group comprising approximately 20% of patients who showed clinical deterioration ^15, 23, 24^. These studies also reported superior predictive performance using GBTM-based classifications, achieving AUCs as high as 0.85 in unadjusted models, compared with 0.66 when using admission NIHSS alone ^15^. In contrast, our study revealed a milder clinical profile overall, with most patients improving rather than deteriorating. While the GBTM-derived categories in our data did improve model fit (AIC 1624 vs. 1759 for admission NIHSS-only model), they provided only modest gains in discrimination (AUC 0.792 vs. 0.778). This suggests that although GBTM can capture subtle data structure and improve internal fit, it is not primarily designed for prediction. By reducing continuous variation into categories, GBTM sacrifices granularity and may introduce residual confounding, meaning that improvements in fit do not necessarily translate into better predictive performance. Hence, previous GBTM-based findings, often drawn from tightly controlled clinical trials, may overstate generalisability and require external validation.

A major strength of our study lies in the use of real-world clinical data to model stroke severity dynamics, enabling an externally valid evaluation of prognostic models compared to those developed using highly selected trial populations. This allowed us to critically review and assess the applicability of previously developed models in routine care settings. Secondly, by anchoring our analyses on time from symptom onset—rather than the more commonly used hospital admission time—we were able to capture early fluctuations in stroke severity that occur before clinical evaluation. This approach reduces potential bias from left truncation, which is frequently overlooked in prior studies and can distort both model estimates and clinical interpretation. Finally, we systematically compared several modelling strategies to evaluate how best to incorporate serial NIHSS assessments into prediction models, offering insights into the most effective ways of leveraging longitudinal severity data to improve prognostic accuracy.

However, the study has limitations. Our sample included only patients with both admission and second NIHSS assessments, excluding a substantial portion of cases with missing values. The assumption that this missingness is completely unrelated to outcome may not hold, limiting the robustness of complete case analysis ^25^. Future work should explore imputation methods under both missing-at-random and missing-not-at-random assumptions to assess the impact on trajectory modelling and performance of prognostic modelling. Additionally, measurement errors in both NIHSS and timing variables may have affected our estimates. The NIHSS, although commonly treated as a continuous and normally distributed outcome, is known to be positively skewed and zero-inflated. Generalised linear mixed models with appropriate distributional assumptions and more flexible latent class regression modelling approach may yield better fit and more robust statistical inference ^26, 27^. Lastly, our models were evaluated using only short-term functional outcomes, which may not reflect long-term recovery patterns.

In conclusion, stroke severity evolves dynamically after symptom onset, and modelling these dynamics, rather than relying on single timepoint measurements or naïve categorisation, improves the statistical efficiency and predictive accuracy of stroke prognostic models. Importantly, these findings highlight the need for incrementally updated prediction models, where new clinical information (such as repeated severity assessments) is incorporated over time. Such dynamic updating reflects how prognosis naturally changes as patient status evolves and underscores the central role of changing stroke severity in outcome prediction. Future prognostic research in this field may consider prioritising advanced longitudinal modelling approaches that not only capture symptom trajectories but also enable updating of predictions in routine clinical settings.

## Statements

## Author contributions

Zewen Lu: Conceptualisation; Methodology; Study design and data analysis; Writing-original draft. Writing-review & editing. Halvor Næss: Data management and acquisition; Ethical approval; Writing-review & editing. Matthew Gittins, Amit K Kishore, Craig J Smith and Andy Vail: Conceptualisation; Methodology; Supervision; Writing-review & editing.

## Conflicting interests

All authors declared no potential conflicts of interest concerning the research, authorship, and/or publication of this article.

## Funding

None

## Ethical considerations

This study was approved by the Regional Ethics Committee for Medical and Health Research Ethics in Western Norway (approval no. 2012/1483 Norstroke).

## Data Availability

All data produced in the present study are available upon reasonable request to the authors.

## Notes

### Competing Interest Statement

The authors have declared no competing interest.

### Funding Statement

This study did not receive any funding

